# Fuzzy logic assisted COVID 19 safety assessment of dental care

**DOI:** 10.1101/2020.06.18.20134841

**Authors:** Andrio Adwibowo

**Affiliations:** U. o. Indonesia., West Java, Indonesia

**Keywords:** dental care, disinfection, fuzzy system, travel history, temperature, ventilation rate

## Abstract

Uncertainty is significant when assessing a risk of certain health care facility conditions especially the facility that prone to the COVID 19 risk. One solution to deal with an uncertainty in health situation assessment is through fuzzy inference system. For that reason, this study aims to develop fuzzy assisted system to assess the safety of dental care related to the sets of patient and environmental conditions. The fuzzy system allows assessment based on the patient’s body temperature, travel history, dental care ventilation rate, and disinfection frequency. The fuzzy system incorporates several steps including fuzzification, fuzzy regulation, and defuzzification. As a result of this study, the fuzzy system is able to assess and identify the risk of dental care according to the patient’s health status and hygiene conditions of dental care as well. To conclude, fuzzy system used in this study has offered the advantage of assessing at any situation as for patient and environmental factor predicts the safety of dental care.

## Introduction

Dental care should be aware on the exposure risks since it is an occupation with a high potential for exposure. On behalf of these exposure risks, all dental care providers must always be cautious and meticulous in mitigating the risk as well as develop and provide clear and easy guidelines and methods to manage early safety measures against any risk (Peditto *et al*. 2020).

Dental care needs improved effective strategies for risk prevention and reduction. Nonetheless, one of the main challenges in the dental care is the difficulty in the various issues including patient and history identification, early warning system and measures, and risk diagnosis. Dental care should ensure that all services are well acquainted to the preventive measures. Provision of dental care should take into consideration the availability of personal protective equipment, ventilations, proper hygiene practice, and routine disinfections (Ahmed *et al*. 2020).

The preventive measures should cover wide aspects including personals, patients, and environments. Dental care should notice a patients with a body temperature measuring >37.5 °C and address a few questions about the patient’s general health status in the last 7 days and history of having been in contact with other infected persons. Personal prevention should be associated with the environmental aspect to prevent the spread of the virus through environmental remediation. This includes to ensure the every surface in the dental care room must be considered at risk. Likewise the dental care environment should provide adequate ventilation rate and all surfaces should be cleaned and sanitized with alcoholic disinfectant (Spagnuolo *et al*. 2020).

Currently, dental care at all times is encouraged to competently follow cross-infection preventive measures (Odeh *et al*. 2020). Especially during this critical time, dental care should have decision making system to decide on the risk and which measures should be taken that are indicated for dental treatment. Dental care should have an update on how this condition is related to their profession in order to be well coordinated, oriented, and prepared. Considering this situation, this study aims to provide approach for a dental care to evaluate input, assess situation, and develop preventive measures using fuzzy systems.

## Methodology

The fuzzy logic rules developed in this study was following Dhiman and Sharma (2020), Muka *et al*. (2017), Princy and Dhenakaran (2016), and Walia *et al*. (2016). The steps are as follows:

### Parameter identification

It is fundamental to first consider all necessary preventive measure parameters that contribute to minimize the risk during visit to dental care. All measures should cover aspects of patients and environments of dental care. Correspondingly, a set of patient and environmental aspects of dental care along with membership function and linguistic ranges were listed in Table 1.

**Table 1.** Parameters and membership values with linguistic ranges.

| Parameters | Membership values with linguistic ranges |
| --- | --- |
| Patient's body temperature | low, medium, high |
| Patient's travel history | low, medium, high |
| Dental care ventilation rate | low, medium, high |
| Dental care disinfection frequency | low, medium, high |

### Membership function (fuzzification)

Membership functions were selected for each value of the input parameters. The degree of an input belongs to a fuzzy set is denoted by membership function ranging from 0 to 1 and expressed as µ. Membership functions were developed for each parameter along with their µ values.

### Fuzzy inference system and rules

The fuzzy inference system to diagnose the dental care risks is developed using 4 input parameters (Table 1) and 1 output as available in Figure 1. Those inputs and outputs were further divided into 3 linguistic ranges denoting the risk levels ranging from low, medium, to high. The “if then” rules were used to analyze the inputs to generate the risk levels. The fuzzy inference system is consisting of 3 steps, including fuzzification or developing the membership function by translating and denoting (µ = 0 – 1) the linguistic ranges, fuzzy regulation or developing the “if then” rules, and defuzzification.

**Figure 1.**
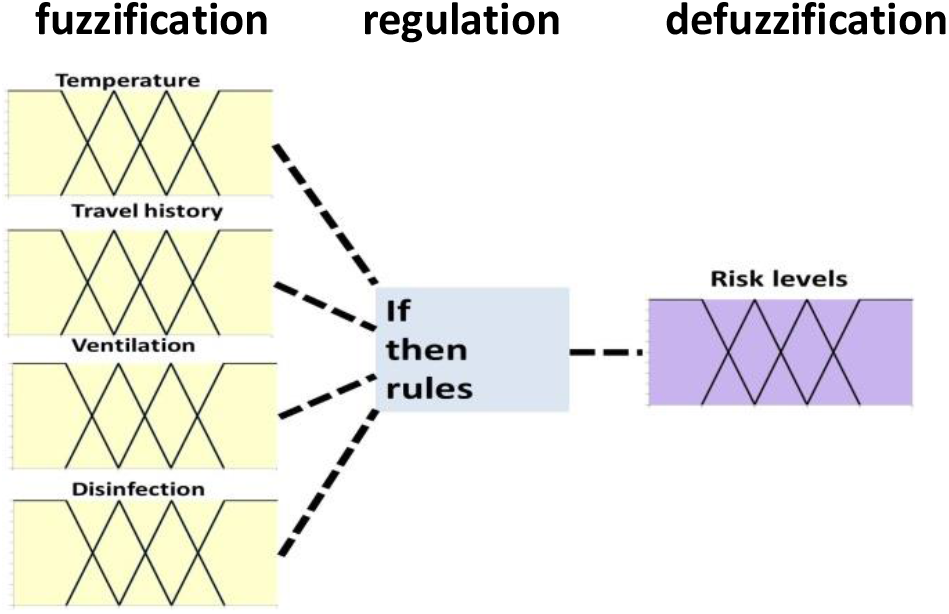
Fuzzy inference system with 4 inputs, 1 output, and if then rules.

## Results

This study has developed 4 input fuzzy set tables. Those fuzzy set tables of membership functions consisted of patient’s body temperature, travelling history, ventilation, and disinfection frequencies. The body temperature input variable supports 4 body temperature levels. This variable was divided into 4 fuzzy sets of linguistic ranges. Fuzzy sets were low, medium, high, and very high. Membership functions of low, medium, high, and very high were triangular. These fuzzy sets and membership functions are shown in Figure 2.

**Figure 2.**
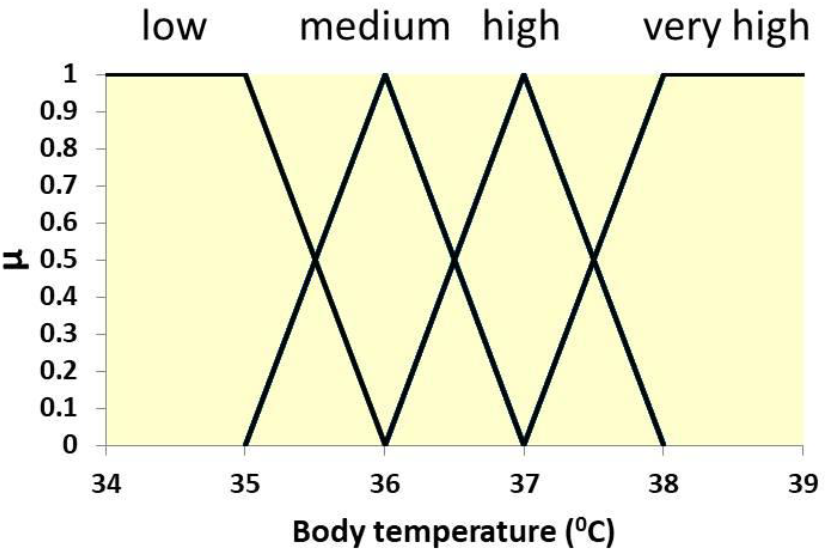
Membership functions of body temperature.

The membership equations and µ values for body temperature for each linguistic type are:

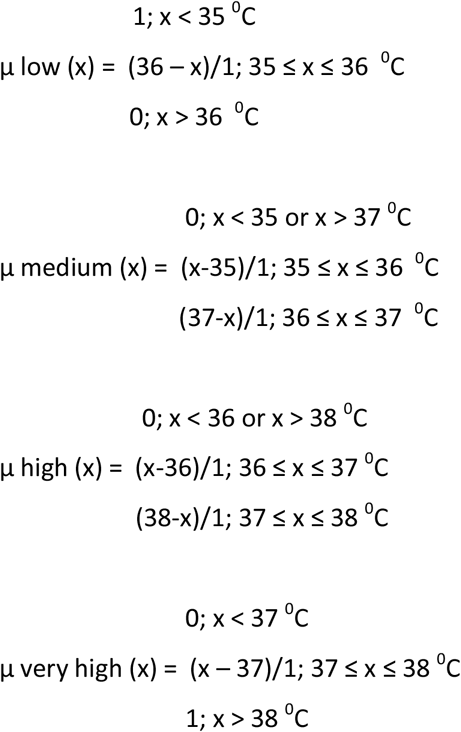

The next variable is related to the travel history of patient in the last 14 days or being in close contact with a person having travel history (Marwaha and Shah 2020). The fuzzy sets of travel history include high, medium, and low risks. High means has travel history within the past 14 days, medium has travel history within the past 14 and 28 days, and low has travel history within the past 28 days prior to visit to dental care (Figure 3).

**Figure 3.**
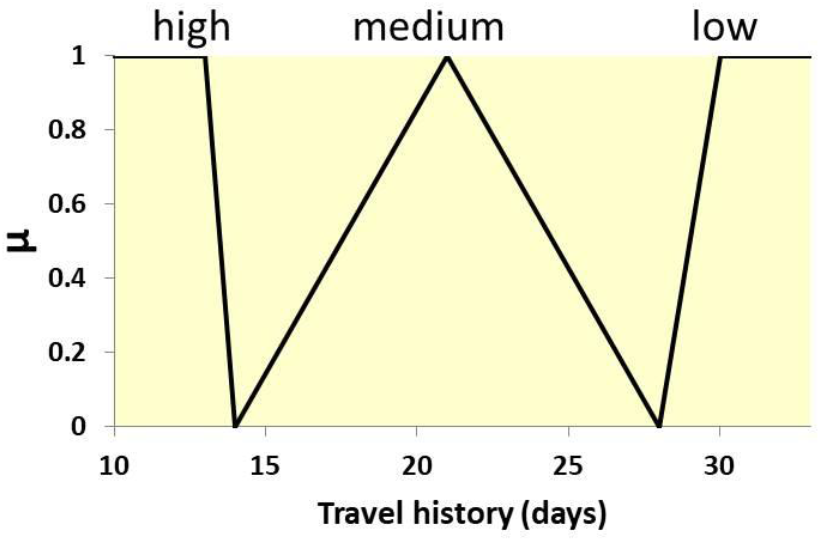
Membership functions of travel history.

The membership equations and µ values for travelling history record for each linguistic type are:

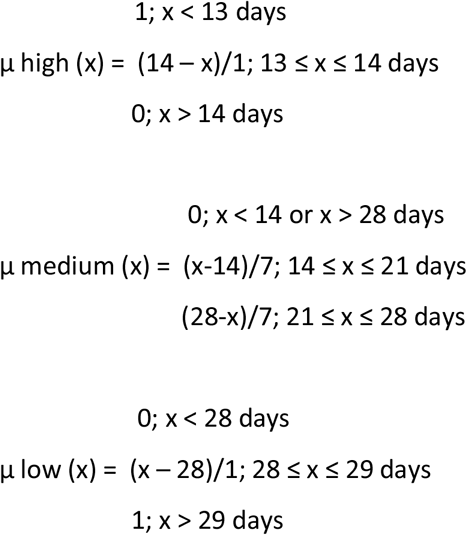

The triangular fuzzy sets for ventilation rates were following data obtained from study by Dai and Zhao (2020). The ventilation rate range of 500 to 800 m^3^/h was defined as the low fuzzy set. While the ranges of medium and high fuzzy sets were 600 to 1000 m^3^/h and 800 to 1100 m^3^/h respectively (Figure 4).

**Figure 4.**
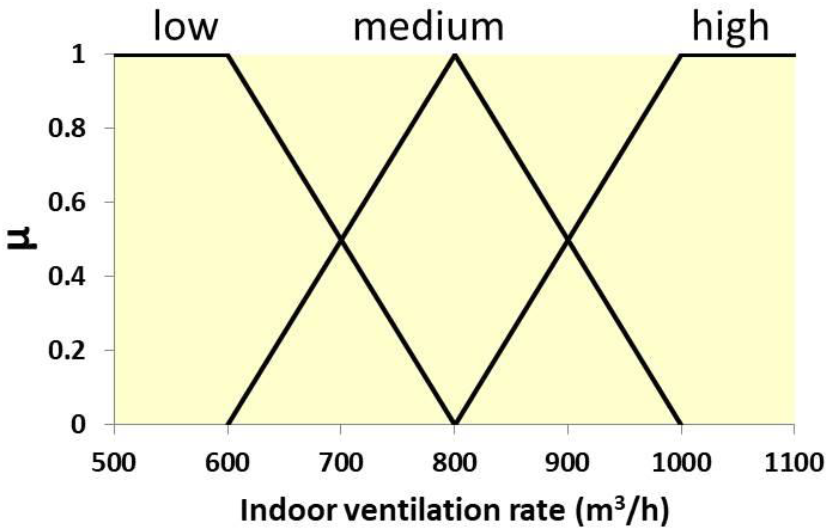
Membership functions of indoor ventilation rate.

Follow is the membership equations and µ values for indoor ventilation rate and linguistic ranges.

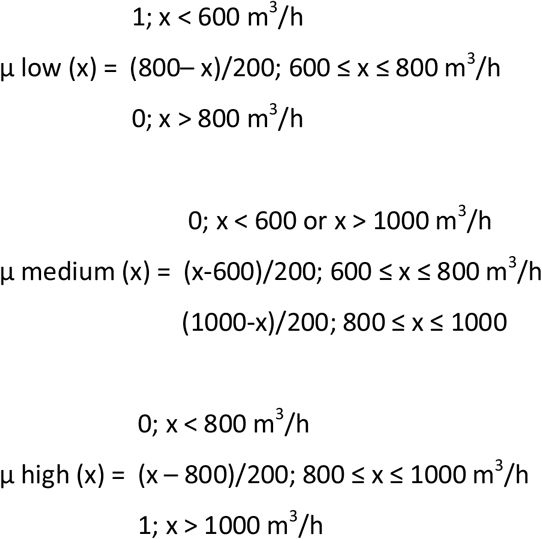

The triangular fuzzy sets for disinfection frequency were divided into low, medium, and high linguistic ranges. The linguistic classification was following how often the dental care was disinfected per day (Figure 5).

**Figure 5.**
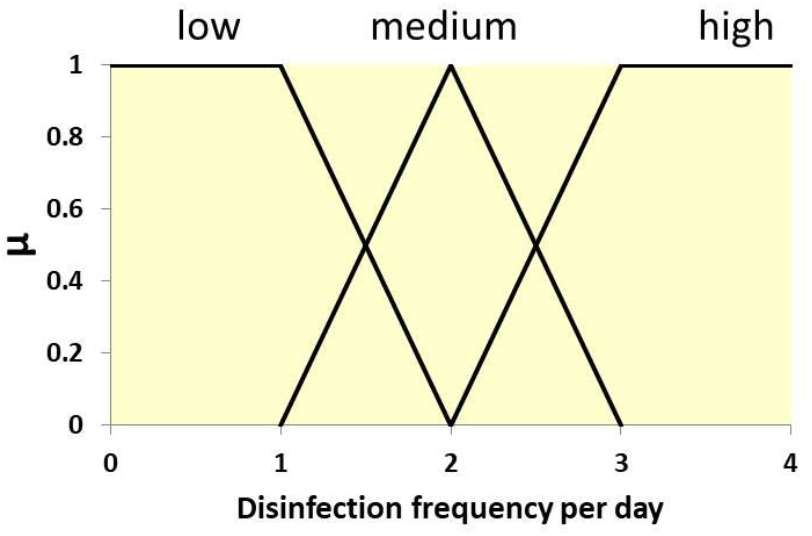
Membership functions of disinfection frequency.

The membership equations and µ values for disinfection frequency for each linguistic type are:

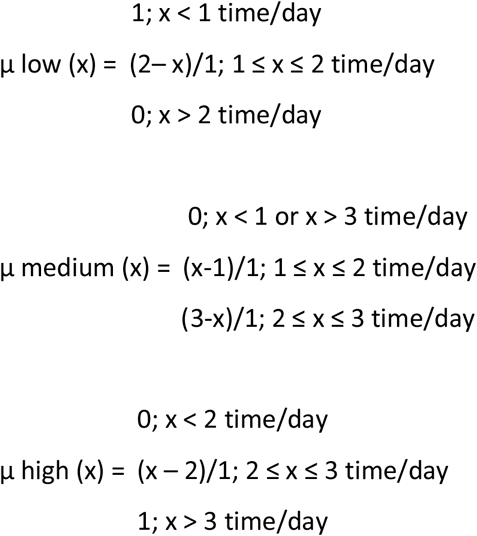

The result of developed fuzzy system in this study was obtained through input of real parameter values (body temperature: 36.9 °C, ventilation rate: 620 m^3^/h), rules, and calculated using defuzzification. Below are the calculation steps.

First define the rules as follows:

1. If patient’s body temperature is high and dental care ventilation rate is low then the risk is high.
2. If patient’s body temperature is medium and dental care ventilation rate is medium then the risk is medium. Calculate the risk of dental care using fuzzification if patient’s body temperature is 36.9 °C and the ventilation rate is 620 m^3^/h.

Second step run the defuzzification:

1. Fuzzification of body temperature membership (Figure 2) at x = 36.9 °C yields µ = 0.4 for medium body temperature fuzzy sets and µ = 0.8 for high body temperature fuzzy sets.
2. While fuzzification of indoor ventilation rate (Figure 4) at x = 620 m^3^/h yields µ = 0.2 for medium ventilation rate fuzzy sets and µ = 0.8 for low ventilation rate fuzzy sets.
3. Based on the rules, the fuzzification values are “If patient’s body temperature is high (µ = 0.8) and dental care ventilation rate is low (µ = 0.8) then the risk is high”, the high risk is equal to minimum µ equal to 0.8.
4. While the fuzzification values for the second rules are µ = 0.4 (body temperature is medium) and µ = 0.2 (dental care ventilation rate is medium), then the medium risk is equal to minimum µ equal to 0.2.
5. The next step is plotting the µ = 0.2 with the risk level of risk fuzzy sets (30) and the µ = 0.8 with the risk level of risk fuzzy sets (90) (Figure 6).
6. The calculation as follows:

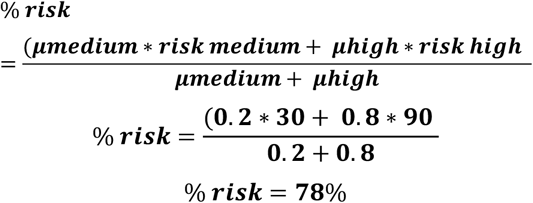

**Figure 6.**
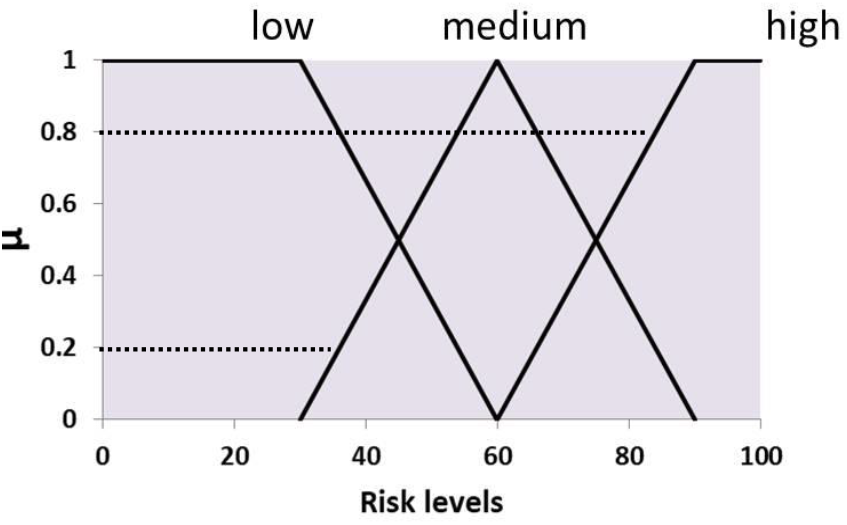
Membership functions of risk levels.

To conclude with patient’s body temperature is 36.9 °C and the ventilation rate sets to 620 m^3^/h, the risk of dental care is 78%. The fuzzy system can assist the dental care to project the risk that the dental care may pose related to the patient’s status and environmental conditions. The risk information can guide the dental care management to make a decision and take a necessary preventive measure.

Sets of rule combinations can be developed based on the available inputs (Table 2). Since this study has recorded 4 input parameters consisting of body temperature, travelling history, ventilation rate, and disinfection frequency and each parameter has 3 linguistic types (low, medium, high), then according to Allahverdi and Ertosun (2018) the numbers of fuzzy rule combinations can be calculated as: 3*3*3*3 = 81 rule combinations

**Table 2.** The fuzzy rule combinations

| Inputs |  |  |  | Risk outputs |
| --- | --- | --- | --- | --- |
| Body temperature | Travelling history | Ventilation rate | Disinfection freq. |  |
| high | high | low | low | high |
| high | high | low | med | high |
| high | high | low | high | high |
| high | high | med | low | high |
| high | high | med | med | high |
| high | high | med | high | high |
| high | med | high | low | high |
| high | med | high | med | high |
| high | med | high | high | high |
| ... | ... | ... | ... |  |
| ... | ... | ... | ... |  |
| ... | ... | ... | ... |  |
| low | low | high | high | low |

## Discussion

The recent pandemic has raised the safety and preventive measures in all health sectors including the face to face dental care. There are a large body of literature has discussed and formulated the necessary dental care safety and preventive measures. Marwaha and Shah (2020) and Iqbal (2020) have identified several important parameters in dental care that should be considered as a part of the safety and preventive measures. Those parameters were including identification, infection control measures, and self protection. The detail of those preventive measures are available in Table 3.

**Table 3.** Safety and preventive measures in linguistic scale of dental care (Marwaha and Shah 2020, Iqbal 2020).

| Parameters | Measures |
| --- | --- |
| Identification | Take proper medical history, body temperature, and travel history (have travelled or have contacted with a person with confirmed 2019-nCoV infection within the past 14 days). |
| Infection control | Personal protection equipment, routine cleaning, and disinfection. |
| Self protection | Use disposable PPE, the PPE should be worn once and discarded after use, and receive flu vaccine. |

Since dental care is provided inside an indoor unit, a hygiene of the indoor room should be considered as well. Some issues regarding the hygiene of dental care are related to the frequencies of disinfections. WHO (2020) recommended that environmental cleaning and disinfection in clinical and health care facilities should follow detailed SOPs. Particular attention should be emphasized on the disinfection of high-touch surfaces and items including light switches, door handles, tables, water/beverage pitchers, and trays and disinfection should be performed routinely and frequently. Especially in the patient rooms, bathrooms, hallways, and corridors the cleaning and disinfection should be at least 3 times daily as recommended.

Regarding the indoor dental care, the ventilation is also an important issue. The ventilation rate did affect the infection probability. The probability was the function of ventilation rate and follow negative correlation. Increases of ventilation rate were observed followed by decrease of infection probability (Dai and Zhao 2020).

Based on those literatures on preventive measures, several measures have been adopted in this study to be calculated using fuzzy logic. Most of the measures are available as linguistic scales. Nonetheless, in this study those linguistic scales have been translated into triangular fuzzy scales for fuzzy system calculations. The translation of linguistic into triangular fuzzy scales aims to deal the ambiguity observed frequently in many fields including in health care. According to Arji *et al*. (2019), most medical concepts are fuzzy, difficult to formalize and measure, and depend upon practice and expertise of healthcare expertize. In medical diagnosis, the meaning of high of blood pressure measurement might pose a different implication depending on the medical history of the patient, the healthcare professional, the clinical context, and this will be resulted in ambiguity (Yunda *et al*. 2015).

Nonetheless fuzzy system is considered can turn that ambiguity observed in medical practice into tangible model to make decision in the middle of inaccuracy, uncertainty, and incompleteness environment. Fuzzy uses a technique to establish accuracy what is oftenly considered vague. Likewise, fuzzy system can make a clear boundary which is before are unclear and vague. By using the fuzzy system, the boundary of elusive symptoms including fevers, fatigue now is clear and tangible.

The fuzzy logics in the field of health care have been widely used and applied in wide ranging of topic from disease diagnosis (Walia *et al*. 2016, Walia *et al*. 2017), ophthalmic ultrasonography retinopathy diagnosis (Andrade *et al*. 2020), and diabetes risk assessment (Allahverdi and Ertosun 2016). Walia *et al*. (2016) estimated that fuzzy set for haemoptysis value of 0.5 and cough 40, the fuzzy system generates output 0.88 which corresponds to a probability of lung infection. The outputs were obtained by employing fuzzy regulation and calculating the input parameters through defuzzification. In diagnosing tuberculosis using fuzzy system, Walia *et al*. (2015) developed several fuzzy regulations as follows:

“if cough less, no chest pain, no haemoptysis, yes loss of appetite, no BCG vaccine, normal fever, less smoke addiction, yes malaise, yes weight loss, then mild tuberculosis”. It can be drawn from that rules that the function of fuzzy system depends on the inputs, rules, and defuzzification.

The COVID 19 related dental care fuzzy system developed in this study is comparable to another COVID 19 fuzzy system. Correspondingly, the latest was the fuzzy inference system developed by Dhiman and Sharma (2020). They have developed a system that able to identify and prevent the risk of COVID 19. They used 11 input parameters including travel history, fever, breathing difficulty, body pain, fatigue, cough, diarrhea, and other medical conditions. Those inputs were translated into 3 linguistic types (low, medium, high). Based on their rules, they inferred that “if the room temperature is medium, body temperature is high, and more intake of ethanol then the severity level of patient is normal”. In contrast, the fuzzy rule used in this study is not deciding the status of patients. Nonetheless, it is illustrating the risk levels or probability of dental care having risks by taking account of patient and environmental status as well.

## Conclusion

Despite there is still a room for improvement, this study has developed a fuzzy system in particular dental care. A patient and environmental status representing the actual conditions of dental care have been represented and calculated in here. The output based on “if then” rules have represented the input parameters conditions. The use of fuzzy system is believed can serve as a dependable and cheap means of diagnosing the risk of dental care may have.

## Recommendation

Dental care like other health cares is facing ambiguity especially when dealing with many elusive health, patient, and environment aspects that require decision making immediately. Correspondingly, a fuzzy system can be used as diagnosis tool for dental care providers to make diagnosis, evaluate and assess the risks, and develop the measures. In future, it is recommended that the fuzzy system incorporates more input parameters to represent all possible risks.

## Data Availability

Data are available in manuscript

